# Circulating SDF4 as a Potential Early Detection Biomarker for Pancreatic Cancer

**DOI:** 10.1101/2025.08.13.25333571

**Authors:** Itsuki Kageyama, Yingsong Lin, Naoki Sasahira, Masato Ozaka, Takashi Sasaki, Makoto Ueno, Hiroshi Ishii, Naoto Egawa, Yasushi Adachi, Tae Sasakabe, Sayo Kawai, Masahiro Nakatochi, Hiroya Yamada, Mitsuro Kanda, Shogo Kikuchi, Asahi Hishida

**Affiliations:** Department of Public Health, Aichi Medical University School of Medicine, Nagakute, Japan; Department of Hepato-Biliary-Pancreatic Medicine, Cancer Institute Hospital, Japanese Foundation for Cancer Research, Tokyo, Japan; Department of Gastroenterology, Kanagawa Cancer Center, Yokohama, Japan; Clinical Research Center, Chiba Cancer Center, Chiba, Japan; Department of Internal Medicine, Tokyo Metropolitan Matsuzawa Hospital, Tokyo, Japan; Department of Internal Medicine, Division of Gastroenterology, Sapporo Shirakaba-dai Hospital, Sapporo, Japan; Department of Gastroenterology and Hepatology, Sapporo Medical University, Sapporo, Japan; Public Health Informatics Unit, Department of Integrated Health Sciences, Nagoya University Graduate School of Medicine, Nagoya, Japan; Department of Hygiene, Fujita Health University School of Medicine, Toyoake, Japan; Department of Gastroenterological Surgery, Nagoya University Graduate School of Medicine, Nagoya, Japan; Aichi Medical University School of Medicine, Nagakute, Japan

**Keywords:** Pancreatic cancer, Early detection, Biomarker, SDF4 (stromal cell–derived factor 4), Blood-based assay

## Abstract

Pancreatic cancer is a highly aggressive malignancy that is often diagnosed at an advanced stage, largely due to the lack of reliable biomarkers for early detection. Stromal cell–derived factor 4 (SDF4), also known as Cab45, is a secreted calcium-binding protein localized to the Golgi apparatus, and has been implicated in various tumor types. However, its clinical relevance in pancreatic cancer remains poorly understood. In this study, we investigated the diagnostic potential of circulating SDF4 levels using data from a hospital-based case–control study in Japan, consisting of 124 patients with pancreatic cancer (stage I–IV) and 125 age- and sex-matched controls. Plasma SDF4 concentrations were measured using an enzyme-linked immunosorbent assay (ELISA). SDF4 levels were significantly elevated in pancreatic cancer patients across all stages compared with controls (p < 0.001). Notably, patients with early-stage diseases (stage I and II) already exhibited substantial increases. Receiver operating characteristic analysis yielded an area under the curve (AUC) of 0.82 for all stages and 0.81 for early-stage patients, indicating good discriminatory performance, including in potentially resectable disease. These findings suggest that SDF4 is elevated as a result of tumor-related biological activity and may represent a promising blood-based biomarker for pancreatic cancer. Its elevation across both early and advanced stage supports further investigation of SDF4 as a complementary marker to carbohydrate antigen 19-9 (CA19-9). Validation in prospective cohorts and in combination with other biomarkers will be essential to clarify its clinical utility.

## Introduction

Pancreatic cancer is a highly aggressive malignancy that is often diagnosed at an advanced stage, resulting in a poor prognosis and a five-year survival rate of only 13%.^1^ In 2022, there were an estimated 511,000 new cases and 467,000 deaths worldwide.^2^ Early detection significantly improves outcomes, with a five-year survival rate of approximately 80% for stage I pancreatic cancer.^3–5^ However, due to its asymptomatic nature in the early stage, less than 10% of cases are diagnosed at this stage. Despite advances in imaging and therapeutic approaches, the lack of effective biomarkers for early-stage detection remains a major clinical challenge.

The most widely used biomarker, carbohydrate antigen 19-9 (CA19-9), lacks sufficient sensitivity and specificity for early detection. Therefore, there is an urgent need for novel, reliable blood-based biomarkers to complement or replace CA19-9 in the early diagnosis of pancreatic cancer. In the search for novel biomarkers, it is important to consider the unique histological and cellular features of pancreatic cancer. One of the hallmark features of pancreatic cancer is its prominent desmoplastic reaction, where stromal cells dynamically interact with cancer cells and secrete a variety of factors into the tumor microenvironment.^6^ Stromal cell–derived factor 4 (SDF4), also known as Cab45, is a secreted calciuml7lbinding protein localized to the Golgi apparatus and involved in the regulation of protein sorting and secretion.^7,8^ In recent years, SDF4 has garnered attention for its elevated expression in multiple tumor types, and emerging evidence suggests its involvement in cancer cell migration, secretion dynamics, and microenvironmental remodeling.^9–11^ Importantly, its expression in both stromal and cancer cells, combined with its ability to be released extracellularly, raises the possibility that SDF4 may enter the circulation and reflect tumor–stromal interactions.^11–13^ These characteristics make SDF4 a compelling candidate for further investigation as a blood-based biomarker in pancreatic cancer.

Although the potential involvement of SDF4 in cancer has been suggested, its clinical utility as a blood-based biomarker for pancreatic cancer remains largely unexplored. In this study, we systematically assessed whether circulating SDF4 levels can serve as a diagnostic marker by analyzing a well-defined cohort of pancreatic cancer patients and matched controls.

## Methods

### Patients

Cases and controls in the present study were selected from a multi-institutional, hospital-based case–control study conducted in Japan, which has been described in detail previously.^14^ For the current analysis, eligible cases were patients aged 20 years or older who were diagnosed with pancreatic cancer at a participating hospital in Tokyo. Diagnoses of invasive pancreatic ductal adenocarcinoma were confirmed by cytology or histopathology, or clinically established based on a poor clinical course consistent with invasive pancreatic ductal adenocarcinoma. Control subjects were cancer-free individuals who underwent medical check-ups at the same hospital. Eligible controls were at least 20 years old with no history of cancer, and were matched to cases based on sex, age, and the date of blood draw. For cases, additional clinical information, including symptoms, histological type, clinical stages based on UICC TNM classifications, and treatment details, was also collected. Blood samples were obtained at the time of enrollment; for patients with pancreatic cancer, samples were obtained between confirmed diagnosis and the initiation of first-line treatment, and stored at –80□°C for subsequent genetic and biomarker analyses. All participants provided written informed consent. Information on demographic characteristics, lifestyle factors (including smoking and alcohol use), and medical history (such as diabetes) was collected using a self-administered questionnaire. This study was approved by the Ethics Committee of Aichi Medical University School of Medicine (Approval No. 2024-M004).

### Enzyme-Linked Immunosorbent Assay

Plasma SDF4 levels were determined using a sandwich enzyme-linked immunosorbent assay (ELISA) kit (abx351752, Abbexa Ltd, Cambridge, UK) following the manufacturer’s protocol. All measurements were performed blinded to clinical information. Most samples were measured in duplicate and the results were averaged; a small number of samples with limited plasma volume were analyzed once. Briefly, 10-fold diluted plasma samples and standards were added to antibody-coated 96-well plates provided with the ELISA kit. After incubation and washing, biotinylated detection antibodies were added to each well. Excess detection antibody was removed by washing, followed by incubation with streptavidin–HRP. After further washing to remove unbound conjugates, a chromogenic substrate solution was added. The reaction was stopped using a stop solution, and absorbance was measured at 450 nm with background correction at 570 nm using an iMark microplate reader (Bio-Rad, Hercules, CA, USA).

### Statistical Analysis

In statistical analyses, comparisons of plasma SDF4 levels between pancreatic cancer patients at different clinical stages and control subjects were first conducted using the Kruskal-Wallis test to evaluate overall differences across multiple groups. When a statistically significant overall effect was observed, post hoc pairwise comparisons were performed using the Wilcoxon rank-sum test with Bonferroni correction to adjust for multiple testing. Pairwise differences between groups were considered statistically significant at a two-sided p-value < 0.05 after correction. Data distributions were inspected visually and assessed for normality prior to selecting nonparametric tests. Receiver operating characteristic (ROC) curve analyses were conducted to evaluate the diagnostic performance of SDF4 in discriminating patients from controls, and the area under the curve (AUC) with 95% confidence intervals was calculated. Statistical significance of each AUC was assessed using a Z-test against the null hypothesis that AUC = 0.5. The optimal cutoff values were determined by maximizing the Youden index (sensitivity + specificity − 1). ROC curve analyses were performed using MedCalc version 20.011 (MedCalc Software Ltd., Ostend, Belgium), and all other statistical analyses were carried out using Stata ver.18 (StataCorp, Collegestation, TX). All reported p-values are two-sided.

## Results

### Clinical characteristics of patients

A total of 249 individuals were included in the analysis: 125 controls and 124 patients with pancreatic cancer (stage I: n = 29, stage II: n = 30, stage III: n = 30, stage IV: n = 35) (Table 1). Controls were matched to cases by age, sex, and blood draw date. All samples were collected at the same hospital using standardized protocols (see Methods for details). No significant differences in age or sex distribution were observed between the groups (p > 0.05).

**Table 1.** Clinical characteristics of controls and pancreatic cancer patients.

| Characteristics | Control<br>(*n* = 125) | Case (All)<br>(*n* = 124) | Stage I<br>(*n* = 29) | Stage II<br>(*n* = 30) | Stage III<br>(*n* = 30) | Stage IV<br>(*n* = 35) |
| --- | --- | --- | --- | --- | --- | --- |
| Age, years | 66.3 ± 8.1 | 66.2 ± 7.9 | 62.9 ± 11.0 | 65.4 ± 7.0 | 68.0 ± 5.3 | 68.0 ± 6.7 |
| Sex, male, *n* (%) | 87 (69.6) | 86 (69.4) | 21 (72.4) | 20 (66.7) | 20 (66.7) | 25 (71.4) |

### Plasma SDF4 levels in pancreatic cancer

Plasma SDF4 levels were significantly elevated in patients with pancreatic cancer compared to controls (Figure 1A). When stratified by disease stage, all stages (I–IV) exhibited significantly higher SDF4 concentrations than controls (p < 0.001 for all comparisons, Wilcoxon’s test with Bonferroni correction). Notably, patients with early-stage disease (stage I and II) already exhibited substantial increases in plasma SDF4 levels compared to controls (Stage I: 4.2 ng/mL [IQR, 2.9-5.8]; Stage II: 4.1 ng/mL [IQR, 3.7-5.3]; Controls: 2.8 ng/mL [IQR, 2.1-3.6]; all p < 0.001), indicating that the increase occurs in the early phases of tumor development (Figure 1B). These findings suggest that SDF4 has potential utility as a biomarker for early detection of pancreatic cancer.

**Figure 1.**
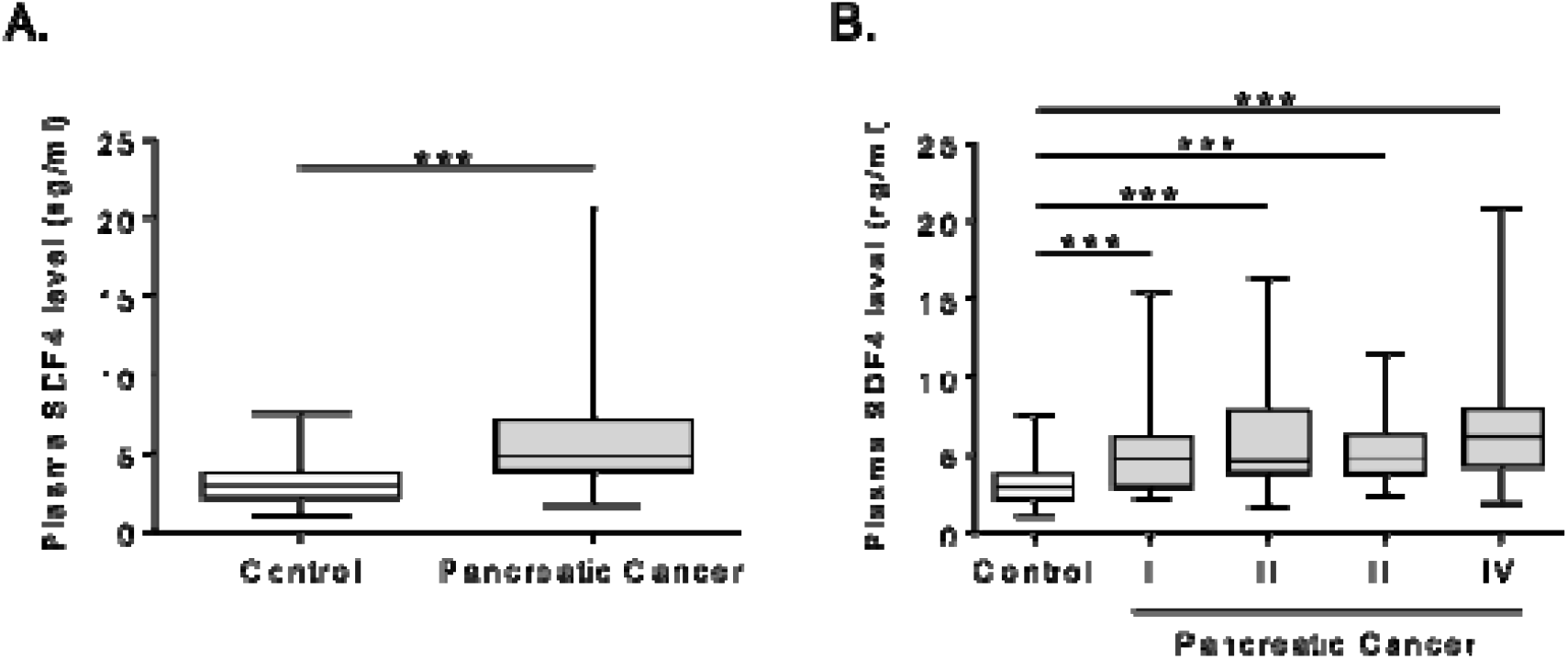
Circulating SDF4 levels in patients with pancreatic cancer and controls. (A) Comparison of plasma SDF4 concentrations between controls (n = 125) and all pancreatic cancer patients (n = 124). (B) Stage-specific distribution of SDF4 levels in patients with stage I (n = 29), stage II (n = 30), stage III (n = 30), and stage IV (n = 35), compared to healthy controls. Statistical analysis was performed using Wilcoxon’s test with Bonferroni correction for multiple comparisons. The box indicates the interquartile range (IQR) from the 25th percentile (Q1) to the 75th percentile (Q3), with the horizontal line inside representing the median (Q2). Whiskers extend to the most extreme data points within 1.5 × IQR from the lower and upper quartiles. ^***^p < 0.001.

### Diagnostic performance of plasma SDF4 in pancreatic cancer

The diagnostic performance of plasma SDF4 was evaluated using ROC curve analysis (Figure 2). For all pancreatic cancer stages (I–IV), the AUC was 0.82 (95% CI: 0.77–0.87), with a sensitivity of 77%, specificity of 78%, and an optimal cutoff value of 3.7 ng/mL. In stage I–II (early stage), the AUC was 0.81 (95% CI: 0.73–0.88), sensitivity 73%, specificity 85%, and cutoff 3.7 ng/mL (Figure 2B). In contrast, stage III–IV (advanced stage) showed an AUC of 0.83 (95% CI: 0.76–0.89), sensitivity 82%, specificity 72%, and cutoff 3.7 ng/mL (Figure 2C). Stage-specific analyses yielded the following results. For stage I, the AUC was 0.85 (95% CI: 0.74–0.93), with a sensitivity of 66%, specificity of 90%, and an optimal cutoff value of 3.6 ng/mL (Figure 2D). In stage II, the AUC was 0.77 (95% CI: 0.65–0.87), sensitivity 80%, specificity 80%, and cutoff 3.7 ng/mL (Figure 2E). Similarly, stage III demonstrated an AUC of 0.79 (95% CI: 0.67–0.89), sensitivity 77%, specificity 77%, and cutoff 3.6 ng/mL (Figure 2F), while stage IV showed an AUC of 0.86 (95% CI: 0.76–0.93), sensitivity 80%, specificity 77%, and cutoff 3.9 ng/mL (Figure 2G). In the analysis of each stage, samples matched for sex, age, and blood draw date were used. All comparisons showed statistically significant discrimination between patients and controls (p < 0.001, DeLong’s test).

**Figure 2.**
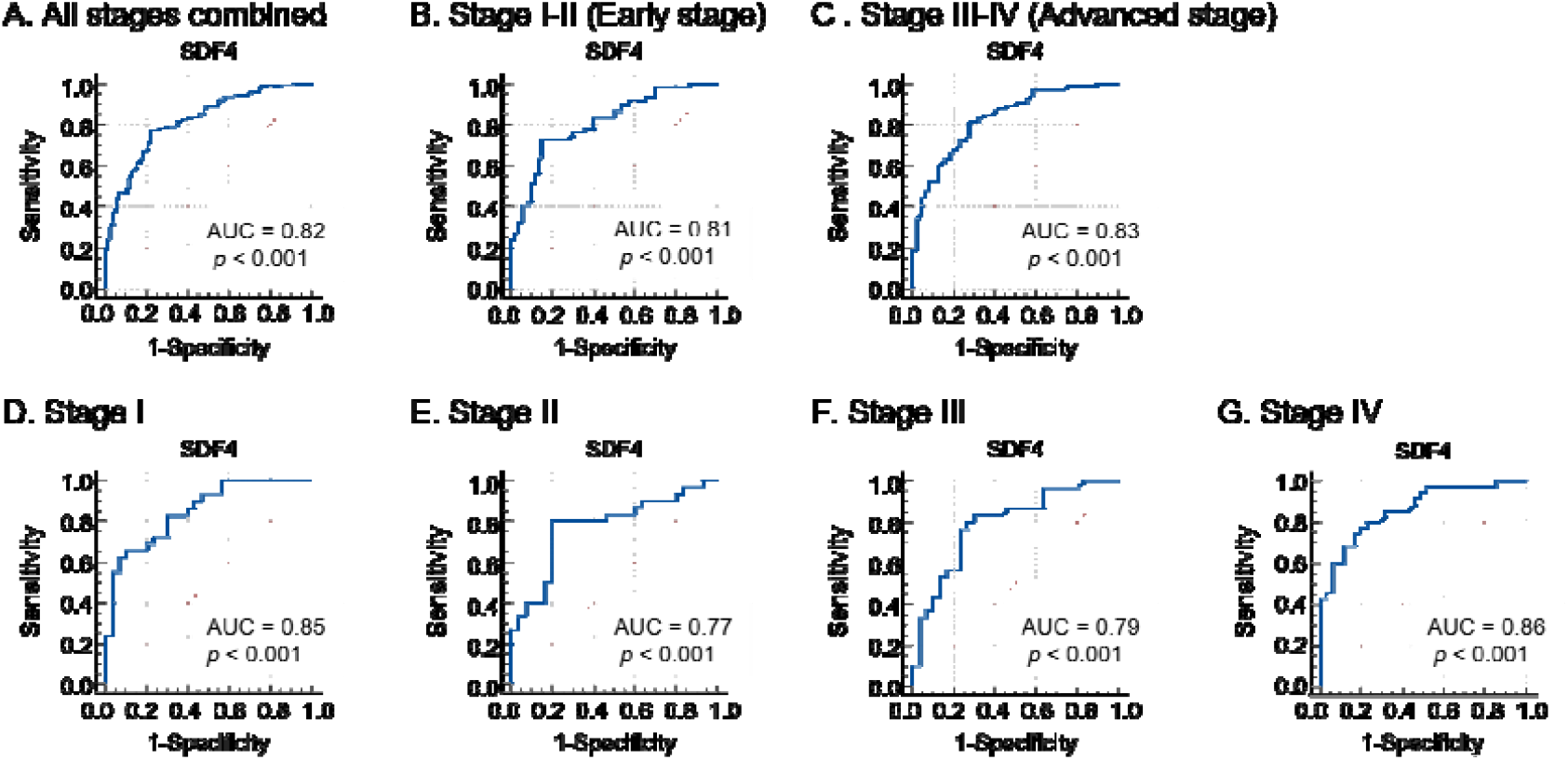
Diagnostic performance of SDF4 for pancreatic cancer detection. (A) ROC curve comparing all stages (I–IV) vs. controls. AUC = 0.82 (95% CI: 0.77–0.87) (B) Stage I–II (Early stage) vs. controls. AUC = 0.81 (95% CI: 0.73–0.88) (C) Stage III–IV (Advanced stage) vs. controls. AUC = 0.83 (95% CI: 0.76–0.89) (D) Stage I vs. controls. AUC = 0.85 (95% CI: 0.74–0.93) (E) Stage II vs. controls. AUC = 0.77 (95% CI: 0.65–0.87) (F) Stage III vs. controls. AUC = 0.79 (95% CI: 0.67–0.89) (G) Stage IV vs. controls. AUC = 0.86 (95% CI: 0.76–0.93) All AUC values were significantly greater than 0.5 (p < 0.001, z-test).

## Discussion

In this study, we found that circulating levels of SDF4 were significantly elevated in patients with pancreatic cancer compared to controls. Blood SDF4 concentration was consistently increased across all disease stages, including stage I-II, and demonstrated strong diagnostic performance in ROC analysis. These findings suggest that SDF4 may be a useful biomarker for the early detection of pancreatic cancer.

While CA19-9 is widely used in clinical practice, its effectiveness for detecting early-stage pancreatic cancer remains suboptimal. Previous studies have shown that CA19-9 may lack sufficient sensitivity for detecting stage I and II pancreatic cancer.^15–17^ Therefore, its use as a sole screeningor diagnostic marker is considered limited. Additionally, CA19-9 levels can be influenced by non-malignant conditions andgenetic factors, including Lewis antigen status.^18,19^ These limitations highlight the need for additional biomarkers that could aid in the early detection of pancreatic cancer. In this study, blood SDF4 concentrations were elevated in all stages of pancreatic cancer, demonstrating good diagnostic performance, particularly in stage I, with an AUC of 0.85 (95% CI: 0.74–0.93). Although a direct comparison was not performed in this study, the consistent elevation of SDF4 across all stages, including early-stage disease, and its diagnostic performance based on ROC analysis suggest that SDF4 may complement existing biomarkers such as CA19-9.

SDF4, also known as Cab45, is a secreted calcium-binding protein localized to the Golgi apparatus.^7,20^ It has been shown to be overexpressed in multiple tumor types, including gastric, breast, colorectal, and pancreatic cancers.^9,10,12,13,21^ Chi et al. (2021) revealed the functional role of SDF4 secreted by cancer-associated fibroblasts in promoting tumor angiogenesis.^11^ A characteristic feature of pancreatic cancer is a marked fibrotic reaction in which stromal cells dynamically interact with cancer cells.^6^ Increased circulating SDF4 levels may reflect the activity of both tumor cells and surrounding fibroblast components. These features suggest that SDF4 is a plausible surrogate marker for tumor–stromal interactions in pancreatic cancer.

Beyond pancreatic cancer, SDF4 was previously identified as a diagnostic candidate in a multi-cancer study.^12^ In that single-institutional analysis, serum SDF4 levels were found to be elevated across several solid tumors, including gastric, colorectal, pancreatic, esophageal, breast, and liver cancers. Notably, the study demonstrated high diagnostic performance for gastric cancer, with an AUC of 0.97, a sensitivity of 89%, and a specificity of 99%, even in early-stage cases. Although the sample sizes for pancreatic and other tumor types were limited, and stage-specific analyses were not conducted for all cancers, the study provided important preliminary evidence supporting the potential utility of SDF4 in a broader oncologic context. In this study, we focused specifically on pancreatic cancer and conducted a detailed evaluation using a well-characterized cohort of 124 patients and 125 matched controls. Through rigorous stage-stratified analyses and a carefully matched case–control design with standardized sample handling, we demonstrated that circulating SDF4 levels were consistently elevated across all disease stages, including early-stage cases. These findings add organ-specific evidence to the growing body of literature on SDF4 and complement prior multi-cancer research^12^, further supporting its potential as a noninvasive diagnostic biomarker in both pancreatic cancer and potentially broader screening applications.

Several limitations of this study should be acknowledged. First, although the sample size is larger than previous reports and includes patients across all stages, it remains modest and is derived from a single institution. External validation in larger, multi-center cohorts is essential to confirm generalizability. Second, although controls were cancer-free at the time of sampling, they were recruited from a medical screening population, and some may have had undiagnosed conditions or developed malignancies after sample collection. This may have introduced some misclassification or heterogeneity in the control group. Third, the study did not include comparisons with benign pancreatic conditions such as chronic pancreatitis or intraductal papillary mucinous neoplasms, which may share biomarker profiles with pancreatic cancer. Whether circulating SDF4 levels are also elevated in chronic pancreatitis, premalignant lesions such as intraductal papillary mucinous neoplasm (IPMN), or chronic diseases including diabetes mellitus remains to be elucidated and warrants further investigation.

In conclusion, our findings indicate that circulating SDF4 is a promising blood-based biomarker for pancreatic cancer. Its consistent elevation across all clinical stages, including early-stage disease, supports its potential utility in early detection. Further studies are warranted to validate these results in larger, multi-center cohorts and to evaluate its performance in combination with existing biomarkers.

## Data Availability

All data produced in the present study are available upon reasonable request to the authors.

## Funding

This work was supported by JSPS KAKENHI Grant Number 23K09646 from the Ministry of Education, Culture, Sports, Science and Technology of Japan.

(All authors have reviewed and approved the final version of the manuscript for submission.)

## References

1. Siegel RL, Kratzer TB, Giaquinto AN, Sung H, Jemal A. Cancer statistics, 2025. CA 269 Cancer J Clin 2025;75:10–45

2. Bray F, Laversanne M, Sung H, Ferlay J, Siegel RL, Soerjomataram I, Jemal A. 271 Global cancer statistics 2022: GLOBOCAN estimates of incidence and mortality 272 worldwide for 36 cancers in 185 countries. CA Cancer J Clin 2024;74:229–63.

3. Blackford AL, Canto MI, Klein AP, Hruban RH, Goggins M. Recent Trends in the 274 Incidence and Survival of Stage 1A Pancreatic Cancer: A Surveillance, Epidemiology, 275 and End Results Analysis. J Natl Cancer Inst 2020;112:1162–9.

4. Klatte DCF, Boekestijn B, Wasser MNJM, Feshtali Shahbazi S, Ibrahim IS, Mieog 277 JSD, Luelmo SAC, Morreau H, Potjer TP, Inderson A, Boonstra JJ, Dekker FW, et al. 278 Pancreatic Cancer Surveillance in Carriers of a Germline CDKN2A Pathogenic 279 Variant: Yield and Outcomes of a 20-Year Prospective Follow-Up. J Clin Oncol 280 2022;40:3267–77.

5. Dbouk M, Katona BW, Brand RE, Chak A, Syngal S, Farrell JJ, Kastrinos F, Stoffel 282 EM, Blackford AL, Rustgi AK, Dudley B, Lee LS, et al. The Multicenter Cancer of 283 Pancreas Screening Study: Impact on Stage and Survival. J Clin Oncol 284 2022;40:3257–66.

6. Erkan M, Hausmann S, Michalski CW, Fingerle AA, Dobritz M, Kleeff J, Friess H. The role of stroma in pancreatic cancer: diagnostic and therapeutic implications. Nat Rev Gastroenterol Hepatol 2012;9:454–67.

7. Scherer PE, Lederkremer GZ, Williams S, Fogliano M, Baldini G, Lodish HF. Cab45, a novel (Ca2+)-binding protein localized to the Golgi lumen. J Cell Biol 1996;133:257–68.

8. Crevenna AH, Blank B, Maiser A, Emin D, Prescher J, Beck G, Kienzle C, Bartnik K, Habermann B, Pakdel M, Leonhardt H, Lamb DC, et al. Secretory cargo sorting by Ca2+-dependent Cab45 oligomerization at the trans-Golgi network. J Cell Biol 2016;213:305–14.

9. Ning J, Liu M, Shen J, Wang D, Gao L, Li H, Cao J. Expression signature and prognostic value of CREC gene family in human colorectal cancer. BMC Cancer 2023;23:878.

10. Luo J, Li Z, Zhu H, Wang C, Zheng W, He Y, Song J, Wang W, Zhou X, Lu X, Zhang S, Chen J. A Novel Role of Cab45-G in Mediating Cell Migration in Cancer Cells. Int J Biol Sci 2016;12:677–87.

11. Chi J-Y, Hsiao Y-W, Liu H-L, Fan X-J, Wan X-B, Liu T-L, Hung S-J, Chen Y-T, Liang H-Y, Wang J-M. Fibroblast CEBPD/SDF4 axis in response to chemotherapy-induced angiogenesis through CXCR4. Cell Death Discov 2021;7:94.

12. Shinozuka T, Kanda M, Shimizu D, Umeda S, Takami H, Inokawa Y, Hattori N, Hayashi M, Tanaka C, Nakayama G, Kodera Y. Identification of stromal cell-derived factor 4 as a liquid biopsy-based diagnostic marker in solid cancers. Sci Rep 2023;13:15540.

13. Grønborg M, Kristiansen TZ, Iwahori A, Chang R, Reddy R, Sato N, Molina H, Jensen ON, Hruban RH, Goggins MG, Maitra A, Pandey A. Biomarker discovery from pancreatic cancer secretome using a differential proteomic approach. Mol Cell Proteomics 2006;5:157–71.

14. Lin Y, Ueda J, Yagyu K, Ishii H, Ueno M, Egawa N, Nakao H, Mori M, Matsuo K, Kikuchi S. Association between variations in the fat mass and obesity-associated gene and pancreatic cancer risk: a case-control study in Japan. BMC Cancer 2013;13:337.

15. Ballehaninna UK, Chamberlain RS. The clinical utility of serum CA 19-9 in the diagnosis, prognosis and management of pancreatic adenocarcinoma: An evidence based appraisal. J Gastrointest Oncol 2012;3:105–19.

16. Hanada K, Shimizu A, Tsushima K, Kobayashi M. Potential of Carbohydrate Antigen 19-9 and Serum Apolipoprotein A2-Isoforms in the Diagnosis of Stage 0 and IA Pancreatic Cancer. Diagnostics (Basel) 2024;14.

17. Ando Y, Dbouk M, Yoshida T, Saba H, Abou Diwan E, Yoshida K, Dbouk A, Blackford AL, Lin M-T, Lennon AM, Burkhart RA, He J, et al. Using Tumor Marker Gene Variants to Improve the Diagnostic Accuracy of DUPAN-2 and Carbohydrate Antigen 19-9 for Pancreatic Cancer. J Clin Oncol 2024;42:2196–206.

18. Liu C, Deng S, Jin K, Gong Y, Cheng H, Fan Z, Qian Y, Huang Q, Ni Q, Luo G, Yu X. Lewis antigen-negative pancreatic cancer: An aggressive subgroup. Int J Oncol 2020;56:900–8.

19. Kim S, Park BK, Seo JH, Choi J, Choi JW, Lee CK, Chung JB, Park Y, Kim DW. Carbohydrate antigen 19-9 elevation without evidence of malignant or pancreatobiliary diseases. Sci Rep 2020;10:8820.

20. Crevenna AH, Blank B, Maiser A, Emin D, Prescher J, Beck G, Kienzle C, Bartnik K, Habermann B, Pakdel M, Leonhardt H, Lamb DC, et al. Secretory cargo sorting by Ca2+-dependent Cab45 oligomerization at the trans-Golgi network. J Cell Biol 2016;213:305–14.

21. Lam PPL, Hyvärinen K, Kauppi M, Cosen-Binker L, Laitinen S, Keränen S, Gaisano HY, Olkkonen VM. A cytosolic splice variant of Cab45 interacts with Munc18b and impacts on amylase secretion by pancreatic acini. Mol Biol Cell 2007;18:2473–80.

